# SystematIc StudY of Patients with atHerosclerotic neUrovascular Stenoses (SISYPHUS): Study Protocol for a Prospective Observational Cohort Evaluating 3D Rotational Angiography and Final Implant Quality in Carotid Stenting

**DOI:** 10.64898/2026.09.20.26363509

**Authors:** Lucas Scotta Cabral, Marco Vugman Wainstein, Sheila Ouriques Martins, Rosane Brondani, Juliana Avila Duarte, Andrea Garcia de Almeida, Angelica Dal Pizzol, Raphael Machado Castilhos, Marco Antonio Stefani, the SISYPHUS Study Executive Committee

## Abstract

**Background and Purpose:** Minor periprocedural ischemic events remain an important limitation of carotid artery stenting (CAS) compared with endarterectomy. Adverse stent–vessel–plaque interactions and in-stent plaque protrusion are proposed as contributing mechanisms. Selective intra-arterial three-dimensional rotational angiography (3DRA) provides high-resolution volumetric imaging directly in the endovascular suite without additional device crossing. Its incremental value for characterizing final implant quality and informing prognosis remains incompletely established.

**Objectives:** We aim to determine whether adding 3DRA to conventional digital subtraction angiography (DSA): (1) improves expert characterization and reproducibility of Final Implant Quality (Imaging Domain); and (2) provides incremental prognostic information for 90-day Textbook Outcome failure beyond baseline clinical risk and conventional DSA (Prognostic Domain).

**Methods:** SISYPHUS is a prospective, multicenter observational cohort of 67 patients undergoing elective CAS across three tertiary centers. Three blinded neuroradiologists will evaluate standardized image packages sequentially under DSA-only and DSA+3DRA conditions. Final Implant Quality will be assessed with a structured 10-item instrument: Q1–Q8 characterize specific findings, Q9 evaluates global implant adequacy (primary imaging variable), and Q10 captures technical image quality. Concealed repeat assessments will be performed in a randomized 17-patient subset for intra-reader reliability. The primary imaging estimand is the within-reader ordinal shift in Q9 after 3DRA is added, modeled with cumulative-link mixed-effects regression. The primary clinical endpoint is 90-day Textbook Outcome failure, integrating early neurological deterioration, major bleeding/procedural complications, stroke, TIA, MI, death, unplanned revascularization, and unplanned readmission within prespecified ascertainment windows. Incremental prognostic performance will be evaluated using nested ridge-penalized logistic regression models (M2 vs M3) with 2,000-resample bootstrap internal validation.

**Trial Status & Prospective Lock:** Recruitment is complete (n=67), while protocol-defined follow-up remains ongoing. The Orthanc/OHIF reader study environment is being implemented; formal reader evaluations have not yet begun. The principal analytical architecture was prospectively locked in the Statistical Analysis Plan (SAP) Version 1.1.1 before formal inspection of reader results and before real-data prognostic model fitting.

## 1. INTRODUCTION

### 1.1 Clinical Background and Problem Statement

Ischemic stroke remains a leading cause of mortality and long-term disability worldwide [1], and extracranial carotid atherosclerosis contributes to approximately 15%–20% of ischemic strokes [2,3]. Carotid endarterectomy (CEA) remains a reference revascularization strategy supported by landmark randomized trials [4–6], while carotid artery stenting (CAS) has become an important alternative in contemporary practice [7,8]. Current decision-making increasingly integrates intensive medical therapy, anatomical suitability, procedural risk, and individualized estimates of future stroke risk.

Despite major advances in technique, embolic protection, antiplatelet therapy, and device design, CAS remains associated with an excess of minor perioperative ischemic events relative to CEA [9–12]. Periprocedural risk is multifactorial and reflects clinical vulnerability, vascular anatomy, plaque biology, procedural factors, and device characteristics [13,14]. Histopathological features of plaque instability are strongly associated with embolic risk, and clinically relevant embolic events may occur after technical completion of the procedure. This interval highlights the need for better characterization of the final treated arterial segment before the patient leaves the angiographic suite.

Beyond conventional major adverse cardiovascular and cerebrovascular events, contemporary carotid cohorts also benefit from multidomain assessment of functional status, cognition, patient-reported outcomes, and unplanned health care utilization. SISYPHUS was therefore designed as a prospective, deeply phenotyped cohort rather than a narrow procedural registry.

### 1.2 Final Implant Quality and the Role of 3D Rotational Angiography

Suboptimal stent–vessel–plaque interaction may contribute to postprocedural embolic risk. Plaque protrusion has historically received particular attention [15], but it represents only one component of **Final Implant Quality**. This broader construct encompasses stent malexpansion, incomplete wall apposition, mesh deformation or fracture, residual plaque near the stent margins, residual luminal stenosis, and mural thrombus. These abnormalities are related but are not assumed to carry equal clinical importance.

Conventional planar DSA is universally available during CAS but may incompletely characterize implant-level abnormalities because of vessel overlap, calcification, and projection dependence. Adjunctive intravascular imaging can reveal abnormalities, including plaque protrusion, that are not fully appreciated on planar angiography [16,17]. However, intravascular imaging requires additional device manipulation across a freshly treated atherosclerotic segment and has not achieved routine use in carotid interventions [17]. Selective intra-arterial 3DRA offers a different strategy. Acquired directly on the angiographic C-arm, it provides submillimeter volumetric information and multiplanar reconstruction without recrossing the implanted stent [18]. Because it can be obtained immediately after standard completion angiography, 3DRA can be studied as an **incremental information layer** added to conventional DSA rather than as a competing stand-alone modality.

### 1.3 Study Rationale and Objectives

SISYPHUS evaluates whether adding 3DRA to completion DSA changes expert characterization of Final Implant Quality and whether that incremental imaging information carries prognostic value for subsequent clinical outcomes. The overall study and analytical architecture is summarized in **Figure 1**.

**Figure 1.**
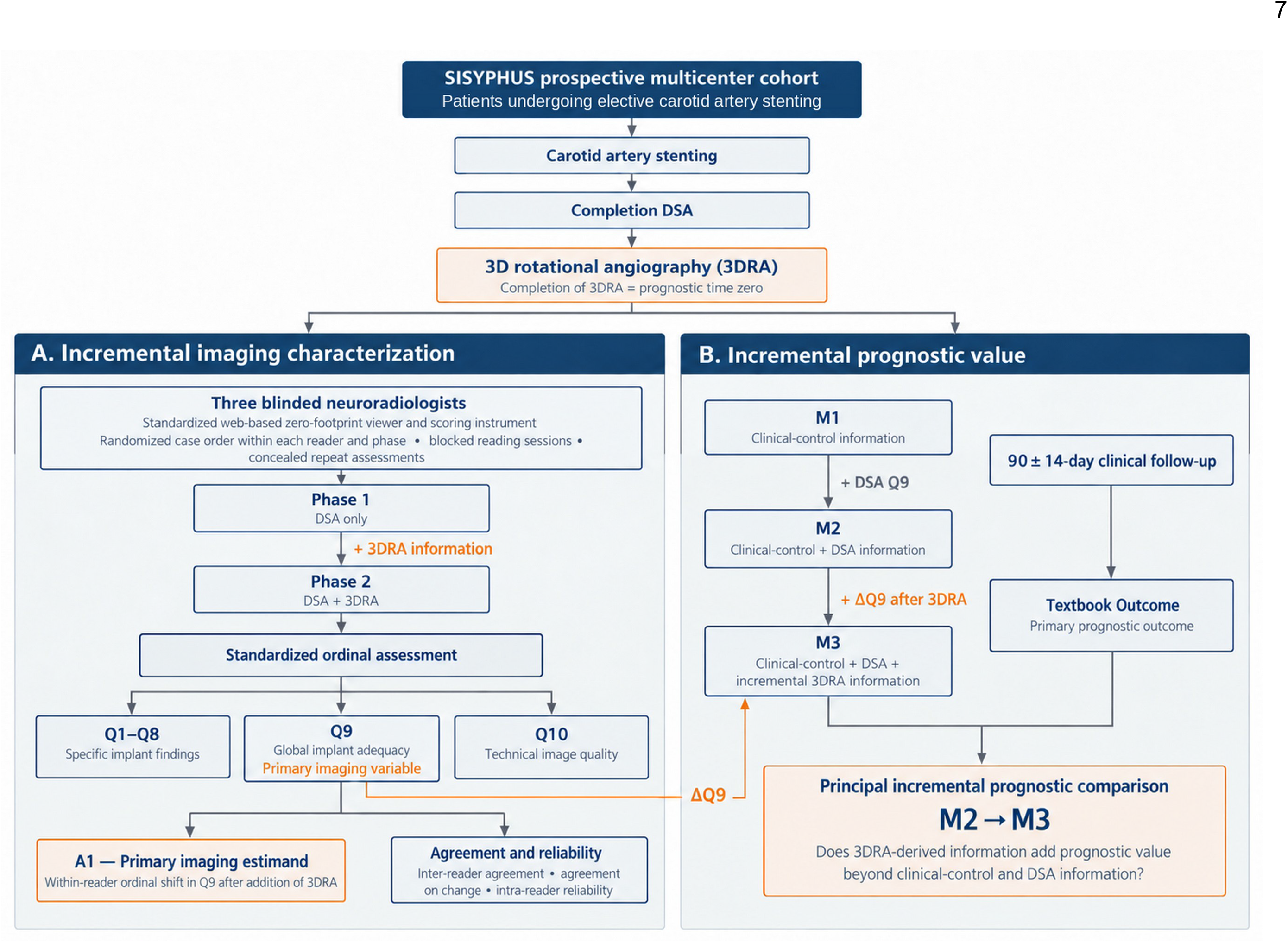
SISYPHUS study design and analytical framework. The figure summarizes the prospective cohort pathway from carotid artery stenting through completion DSA and 3DRA, the sequential blinded reader study, derivation of the primary imaging estimand (within-reader Q9 shift), and the nested M1→M2→M3 prognostic framework used to evaluate the incremental prognostic value of 3DRA-derived information.

The **Imaging Domain** asks whether additional 3DRA information produces a systematic within-reader change in the integrated assessment of implant adequacy and whether the resulting information is reproducible across and within readers. The study does not treat either DSA or 3DRA as an anatomical gold standard for the complete Final Implant Quality construct; the primary estimand is the incremental change in expert assessment.

The **Prognostic Domain** asks whether the information added by 3DRA improves prediction of 90-day Textbook Outcome failure beyond prespecified clinical-control information and conventional DSA assessment. We hypothesize that 3DRA will alter expert characterization of Final Implant Quality and provide incremental prognostic information. Analytical estimands, reader procedures, and model families were prospectively specified before formal image-reader evaluation.

## 2. Materials and Methods

### 2.1 Study Design, Setting, and Ethical Framework

SISYPHUS is a prospective, multicenter observational cohort of patients undergoing elective CAS at three tertiary referral centers in Porto Alegre, Brazil: Hospital de Clínicas de Porto Alegre (HCPA; coordinating center), Hospital Moinhos de Vento, and Hospital Mãe de Deus. The cohort addresses two linked questions: incremental imaging characterization of the final carotid implant and the prognostic value of the information added by 3DRA.

The protocol was approved by the HCPA Research Ethics Committee (CAAE 76550023.4.3002.5330) and prospectively registered in the Registro Brasileiro de Ensaios Clínicos (REBEC: RBR-6cs4wcc). All participants or legally authorized representatives provided written informed consent before study procedures. The study was conducted under applicable Brazilian ethical, institutional, and data-protection requirements [19] and was aligned with Good Clinical Practice principles [20].

The principal analytical architecture was prospectively specified in SAP Version 1.1.1 before formal inspection of reader results and before real-data prognostic model fitting. The SAP structure was informed by the Gamble framework [21] and its observational-study adaptation [22]. Reporting is aligned with STROBE [23], SAMPL [24], GRRAS [25], and relevant TRIPOD+AI principles [26]. The study evaluates incremental prognostic information and does not claim development or external validation of a new clinical risk score.

### 2.2 Study Population, Eligibility Criteria, and Screening

The final prospective cohort comprises 67 participants recruited from Q1 2024 through Q3 2026. No retrospective extension cohort or historical clinical cases will be added to the primary analytical population. Screening logs prospectively documented all potentially eligible candidates and reasons for non-enrollment; the primary results report will include a STROBE flow diagram.

The original protocol anticipated approximately 210–231 participants based on expected clinical-event rates. A smaller-than-planned number of active interventionalists, major disruption of regional health care and research operations during the May 2024 Rio Grande do Sul floods, and the fixed academic timeline of the doctoral program curtailed recruitment. Recruitment was closed without interim inspection of study outcomes and without outcome-driven sample-size re-estimation. The study therefore does not claim preservation of the originally planned precision for rare clinical events and will not use retrospective power calculations to reinterpret findings. Inference will instead emphasize effect estimates, confidence intervals, and internal-validation diagnostics.

*Inclusion Criteria*

1. Multidisciplinary clinical indication for elective CAS scheduled within 30 days.
2. Atherosclerotic carotid stenosis extending from the mid-terminal common carotid artery to the C2 internal carotid artery, in symptomatic or asymptomatic patients.
3. Age ≥18 years.
4. Consent and ability to complete structured follow-up through 90 days.

*Exclusion Criteria*

1. Emergency CAS for acute ischemic stroke or urgent revascularization.
2. Non-atherosclerotic carotid disease (eg, dissection or fibromuscular dysplasia).
3. Retreatment of a previously stented carotid segment.
4. Planned contralateral carotid intervention or major cardiovascular surgery within 30 days.
5. Carotid endovascular thrombectomy before stent deployment.
6. No documented negative pregnancy test when applicable.
7. Severe iodinated-contrast allergy not manageable with standard premedication.
8. End-stage renal disease approaching dialysis with nephrological contraindication to iodinated contrast.
9. Severe hemorrhagic diathesis contraindicating dual antiplatelet therapy.
10. Inability or clinical unsuitability to adhere to prescribed dual antiplatelet therapy.
11. Severe baseline neurological sequelae precluding reliable ascertainment of new events.
12. Severe systemic disease with life expectancy <12 months or advanced palliative-care directives.

Emergency CAS during hyperacute stroke was excluded prospectively because neurological trajectories in that setting are dominated by the presenting ischemic injury and because emergency workflows preclude standardized baseline phenotyping and end-of-procedure 3DRA acquisition.

### 2.3 Revascularization Decision-Making

CAS indication, timing, and modality selection were determined by a multidisciplinary neurovascular team including stroke neurologists, vascular surgeons, and interventional neuroradiologists, consistent with contemporary guideline-based care [11,12]. Symptomatic disease followed established evidence-based thresholds. For asymptomatic disease, luminal stenosis ≥60% served as an initial benchmark rather than an isolated indication. Decisions integrated estimated stroke risk under medical therapy, plaque-vulnerability features, life expectancy, anatomical suitability, expected procedural risk, and patient preference. This approach was intended to reflect real-world multidisciplinary selection rather than enrollment based solely on percentage stenosis.

### 2.4 Baseline Evaluation and Longitudinal Phenotyping

Baseline evaluation was multidimensional. Vascular imaging with CTA and/or MRA characterized stenosis severity, plaque morphology, lesion length, contralateral carotid occlusion, tandem disease, and aortic-arch anatomy. Brain CT and/or MRI assessed pre-existing ischemic injury and white-matter disease, including ASPECTS [27] and Fazekas grading [28]. Duplex ultrasonography was used as an adjunct when needed. Baseline symptomatic status was defined by an ipsilateral ischemic event within the preceding 6 months.

Clinical risk and functional reserve were characterized using ASA physical status [29], the Revised Cardiac Risk Index [30], Duke Activity Status Index [31], the Wimmer/SAPPHIRE Worldwide CAS risk score [32], the Siena CAS score [33], the Oxford/ECST Carotid Stenosis Risk Prediction Tool [34], and PRECISE-DAPT [35]. These scores were recorded as prespecified descriptors or benchmark constructs and were not used to drive variable selection in the primary prognostic model.

Neurological and functional status were assessed with NIHSS [36] and the modified Rankin Scale, including the simplified modified Rankin Scale questionnaire for structured follow-up [37]; frailty with the Clinical Frailty Scale [38], with CFS ≥5 prespecified as the primary binary frailty representation; cognition with the Brazilian-Portuguese TICS-M [39]; health-related quality of life with EQ-5D-3L scored using a Brazilian value set [40]; and depressive symptoms with PHQ-9 and PHQ-2 [41,42]. Standard laboratory assessment included hematological, renal, metabolic, inflammatory, and cardiac biomarkers. Cardiac evaluation included 12-lead ECG and transthoracic echocardiography. **Table 1** summarizes the timing of repeated clinical, laboratory, imaging, cognitive, and patient-reported assessments.

**Table 1.**
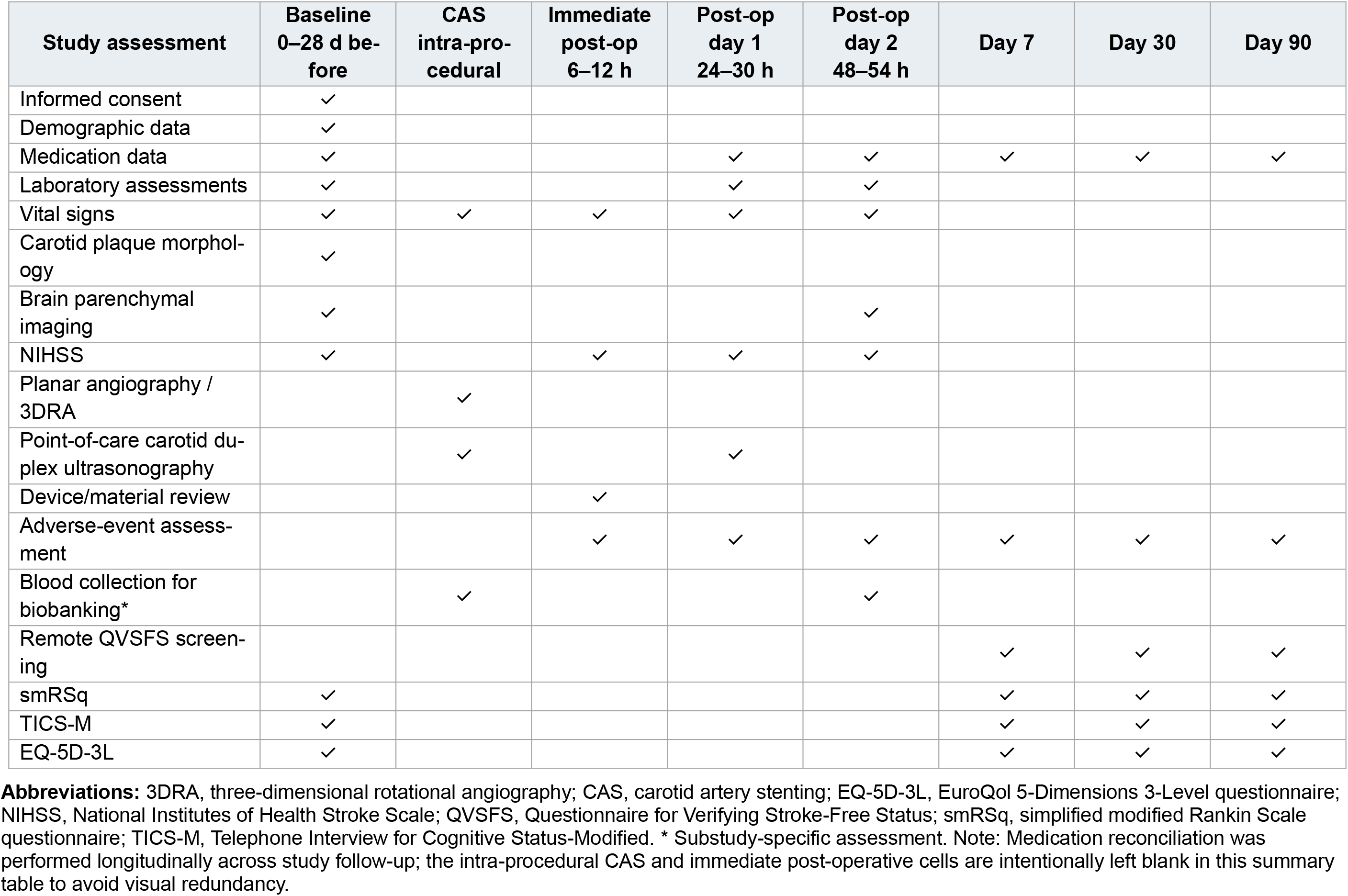
Schedule of Study Assessments and Data Collection.

| Study assessment | Baseline<br>0–28 d before | CAS<br>intra-pro-<br>cedural | Immediate<br>post-op<br>6–12 h | Post-op<br>day 1<br>24–30 h | Post-op<br>day 2<br>48–54 h | Day 7 | Day 30 | Day 90 |
| --- | --- | --- | --- | --- | --- | --- | --- | --- |
| Informed consent | ✓ |  |  |  |  |  |  |  |
| Demographic data | ✓ |  |  |  |  |  |  |  |
| Medication data | ✓ |  |  | ✓ | ✓ | ✓ | ✓ | ✓ |
| Laboratory assessments | ✓ |  |  | ✓ | ✓ |  |  |  |
| Vital signs | ✓ | ✓ | ✓ | ✓ | ✓ |  |  |  |
| Carotid plaque morphol-<br>ogy | ✓ |  |  |  |  |  |  |  |
| Brain parenchymal<br>imaging | ✓ |  |  |  | ✓ |  |  |  |
| NIHSS | ✓ |  | ✓ | ✓ | ✓ |  |  |  |
| Planar angiography /<br>3DRA |  | ✓ |  |  |  |  |  |  |
| Point-of-care carotid du-<br>plex ultrasonography |  | ✓ |  | ✓ |  |  |  |  |
| Device/material review |  |  | ✓ |  |  |  |  |  |
| Adverse-event assess-<br>ment |  |  | ✓ | ✓ | ✓ | ✓ | ✓ | ✓ |
| Blood collection for<br>biobanking* |  | ✓ |  |  | ✓ |  |  |  |
| Remote QVSFS screen-<br>ing |  |  |  |  |  | ✓ | ✓ | ✓ |
| smRSq | ✓ |  |  |  |  | ✓ | ✓ | ✓ |
| TICS-M | ✓ |  |  |  |  | ✓ | ✓ | ✓ |
| EQ-5D-3L | ✓ |  |  |  |  | ✓ | ✓ | ✓ |

**Abbreviations:** 3DRA, three-dimensional rotational angiography; CAS, carotid artery stenting; EQ-5D-3L, EuroQoL 5-Dimensions 3-Level questionnaire; NIHSS, National Institutes of Health Stroke Scale; QVSFS, Questionnaire for Verifying Stroke-Free Status; smRSq, simplified modified Rankin Scale questionnaire; TICS-M, Telephone Interview for Cognitive Status-Modified. \* Substudy-specific assessment. Note: Medication reconciliation was performed longitudinally across study follow-up; the intra-procedural CAS and immediate post-operative cells are intentionally left blank in this summary table to avoid visual redundancy.

### 2.5 Endovascular Procedure and End-of-Procedure Imaging

#### 2.5.1 Carotid Artery Stenting

Periprocedural care followed a standardized CAS pathway. Dual antiplatelet therapy with aspirin and a P2Y12 inhibitor and high-intensity statin therapy were prescribed, with clopidogrel loading when indicated [43]. Procedures were performed under anesthesiologist-supervised conscious sedation with local anesthesia and continuous invasive arterial pressure monitoring. Ultrasound guidance was preferred for arterial access, and the transradial route was favored when anatomically feasible.

Unfractionated heparin (80 IU/kg) was administered before arch navigation, with a target activated clotting time of approximately 2.0–2.5 times baseline. Distal filter embolic protection was routinely used. Stent choice was individualized to plaque morphology and vessel diameter, while device architecture was prospectively captured as a study variable. Post-dilation was selective. Technical success required adequate lesion coverage, ≤30% residual stenosis, and absence of visible distal embolization.

Hemodynamic responses, vasopressor or atropine requirements, procedure times, and technical complications—including access-site injury, dissection, vasospasm, filter complications, arrhythmia, and persistent hypotension/bradycardia—were prospectively recorded. These downstream procedural phenomena are not part of the primary baseline clinical-control model because they occur after the prognostic time origin.

#### 2.5.2 Completion DSA and 3DRA

At the end of CAS, completion DSA was acquired in at least two orthogonal projections centered on the treated carotid segment, with both subtracted and non-subtracted runs retained. This conventional DSA represents the baseline post-stenting imaging information state for the reader study.

Immediately after completion DSA, selective intra-arterial 3DRA was acquired using a standardized single-phase rotational protocol. The access catheter was positioned proximal to the stent, and participants were instructed to minimize motion and swallowing during the sweep. Ten milliliters of nonionic iodinated contrast were diluted with normal saline to a total injectate volume of 50 mL (20% contrast concentration) and injected synchronously with the rotational acquisition. The resulting reconstructions provided submillimeter volumetric information without crossing the freshly implanted stent with an additional intravascular imaging device.

To minimize co-intervention bias, participating interventionalists prospectively agreed not to use the investigational 3DRA reconstructions to modify immediate intra-procedural or postprocedural management. Completion of 3DRA defines prognostic time zero. **Supplement 2** provides detailed acquisition parameters, image-package preparation, and reader study implementation.

### 2.6 Standardized Post-Procedural Care

After CAS, participants underwent structured neurological and hemodynamic surveillance in a neurocritical care or specialized stroke unit. The general systolic blood pressure target was 90–160 mm Hg, with tighter peri-revascularization control when clinically indicated to reduce both hypoperfusion and cerebral hyperperfusion risk. Vasoactive therapy was used as needed under intensive-care supervision. Dual antiplatelet therapy was continued for 30–90 days according to stent design and individual vascular risk, followed by long-term single antiplatelet therapy; high-intensity statin therapy was continued.

Point-of-care duplex ultrasonography at 18–24 hours assessed carotid stent patency/flow and the arterial access site. Procedure-related complications, neurological changes, ICU/ward transitions, and discharge timing were prospectively recorded using standardized study documentation. Follow-up assessments and collections are summarized in **Table 1**.

### 2.7 Reader Study Protocol

Three board-certified interventional neuroradiologists, each with >5 years of post-specialization experience, will evaluate the image packages. Readers will be blinded to baseline clinical information and clinical outcomes and will not participate in endpoint adjudication. Each reader will complete two sequential, non-interleaved information conditions: **Phase 1, DSA only**, followed by **Phase 2, DSA+3DRA**. All Phase 1 cases will be completed before Phase 2 begins, with a washout interval of at least 1 week.

The fixed DSA→DSA+3DRA sequence reflects the scientific estimand: what changes when additional 3DRA information is made available after the baseline DSA assessment. Reversing modality order would contaminate the DSA-only state; accordingly, phase and presentation-order effects are intrinsically non-separable. Potential recall and learning effects will be mitigated through washout, randomized case order within each reader and phase, blocked reading sessions, concealed repeats, and standardized pre-reading training.

Coded DICOM packages will be reviewed in a standardized web-based, zero-footprint Orthanc 1.12.3/OHIF 3.8.0 environment using a prespecified set of viewing interactions. Each case will be evaluated in a single scheduled presentation, and final Q1–Q10 ratings will be locked after submission. The reader study will comprise **402 first-pass assessments** (67 patients × 3 readers × 2 phases). A reproducibly randomized subset of 17 patients (25.4%) will be re-presented covertly within each phase to each reader, contributing 102 repeat assessments and **504 total readings**. Concealed repeats will be used for intra-reader reliability and will not add observations to the primary phase comparison.

Case order will be randomized independently for every reader and phase, and repeat presentations will be placed in nonadjacent blocks. Before formal readings, readers will complete standardized training using 4–5 historical non-study cases to familiarize themselves with the viewer, permitted tools, and scoring instrument without calibration toward a consensus answer. Full operational details, seeds, package restrictions, blocking, repeat implementation, and analysis populations are provided in **Supplement 2**.

### 2.8 Final Implant Quality Assessment

Final Implant Quality is defined as the integrated anatomical and mechanical characterization of the interaction between the deployed stent, arterial wall, atheromatous plaque, and residual lumen immediately after CAS. The construct is assessed using a study-developed 10-item ordinal instrument.

**Q1–Q8** assess specific findings: stent malexpansion; significant mesh deformation or fracture; incomplete stent-wall apposition; eccentric plaque relative to the stent mesh; residual plaque beyond or within 5 mm of a stent margin; residual in-stent stenosis >30%; mural thrombus; and in-stent plaque protrusion. Each is rated on a 5-category scale from definitely absent to definitely present. These findings are not assumed to have equal clinical importance, and no simple unweighted additive Q1–Q8 score is used as the primary quality measure.

**Q9, Global Implant Adequacy**, is the primary integrated imaging variable. It records the reader’s confidence that the implant has been completed successfully and satisfactorily on a 5-category ordinal scale: definitely inadequate, probably inadequate, equivocal/indeterminate, probably adequate, or definitely adequate.

**Q10** records technical image quality as poor/inadequate, acceptable, or excellent and is analyzed separately as a quality-control variable rather than as part of the Final Implant Quality construct. The complete reader-facing instrument, response anchors, English publication translation, and original Brazilian Portuguese wording are provided in **Supplement 1**.

### 2.9 Follow-Up, Clinical Outcomes, and Adjudication

Prognostic time zero is the exact completion of 3DRA acquisition. Participants underwent structured inpatient assessment after CAS and are followed longitudinally through 90 ± 14 days; day 104 is the administrative upper limit for outcome components without shorter prespecified windows. During the index admission, neurological examinations and clinically indicated imaging were used to capture early deterioration and complications. Outpatient surveillance at days 7, 30, and 90 includes medication and adverse-event review, functional/cognitive and patient-reported assessments, and structured screening for recurrent cerebrovascular symptoms with the QVSFS [44]. Positive screens trigger clinical and neuroimaging assessment by a stroke neurologist. The complete assessment schedule is shown in **Table 1**.

Clinical surveillance includes death, stroke, TIA, MI, unplanned revascularization, emergency-department returns, hospital readmissions, and other prespecified adverse events. Short-window index complications are distinguished from later follow-up events: END is ascertained only within 48 hours after time zero, while BARC ≥3 bleeding and Clavien-Dindo ≥3 procedural/surgical complications are ascertained through discharge, up to a maximum of 72 hours. Closure of these early windows does not terminate observation for the remaining outcome components.

#### Primary Clinical Endpoint — Textbook Outcome Failure

Textbook Outcome (TO) is a multidomain patient-centered composite intended to represent an uncomplicated postprocedural trajectory rather than a narrow major adverse event endpoint. It incorporates neurological injury, major procedural harm, treatment failure, and unplanned health care utilization. For prognostic modeling, the event is TO Failure (Y=1); successful TO is coded Y=0.

TO Failure occurs if any of the following is observed within its prespecified ascertainment window:

**1. Early Neurological Deterioration (END)**: NIHSS increase ≥4 within 48 hours of time zero.
**2. Major bleeding**: BARC grade ≥3 [45] within 72 hours.
3. **Major surgical/procedural complication**: Clavien-Dindo grade ≥3 [46] within 72 hours; intracranial hemorrhage is classified according to ECASS-III [47].
**4. Stroke** through day 104.
**5. Transient ischemic attack** through day 104.
**6. Myocardial infarction** through day 104.
**7. All-cause mortality** through day 104.
**8. Any additional unplanned revascularization** through day 104.
**9. Any unplanned readmission**, defined as an unplanned emergency-department return and/or hospital readmission, through day 104.

The inclusion of unplanned utilization was prespecified because a clinically meaningful “textbook” recovery requires not only freedom from major organ injury but also freedom from treatment failure and unexpected return to acute care. The binary TO endpoint remains the primary prognostic outcome; prioritized time-to-event and win-statistic decompositions are complementary analyses rather than alternative primary endpoints.

#### Secondary Clinical Composite — Adverse Outcome

The secondary **Adverse Outcome** composite is intentionally narrower than TO. It focuses on canonical cardiovascular and cerebrovascular events and failure of the initial revascularization strategy: stroke, TIA, MI, all-cause death, or any additional unplanned revascularization during follow-up. END, index-admission bleeding/procedural complications, and readmission are excluded from this composite. Individual components are also evaluated separately.

#### Independent Endpoint Adjudication

All clinical outcomes and adverse-event records will undergo independent review by the study Clinical Endpoint Adjudication Committee. Adjudicators will be blinded to image-reader findings and will have access to the relevant clinical records, laboratory data, and diagnostic/neuroimaging studies required for standardized event classification. The committee will issue the final event classification.

### 2.10 Ancillary Substudies

The cohort supports four prespecified ancillary domains that broaden phenotypic characterization without redefining the primary objectives: (1) continuous bilateral transcranial Doppler monitoring during CAS for intra-procedural high-intensity transient signals; (2) blood-based brain-injury/inflammatory biomarkers with cryopreserved biobank samples; (3) longitudinal cognitive and functional assessment, including TICS-M; and (4) patient-reported health-related quality of life with EQ-5D-3L and depressive-symptom assessment. These data are intended for dedicated secondary/substudy analyses.

### 2.11 Statistical Analysis

#### 2.11.1 General Principles

Analyses will follow SAP Version 1.1.1. Tests will be two-sided with α=.05 and 95% confidence intervals, with interpretation emphasizing effect magnitude, direction, precision, calibration, consistency, and clinical plausibility rather than isolated significance thresholds. Baseline characteristics will be summarized descriptively, and primary predictors will not be selected on the basis of univariable significance. No stepwise selection will be used in the primary prognostic models.

Multiplicity will be addressed principally through a prespecified hierarchy rather than a family-wise confirmatory correction across every analysis: one **primary imaging estimand (A1)** and one **principal incremental prognostic comparison (M2 vs M3)**. Secondary, sensitivity, and exploratory findings will not independently redefine the primary study conclusion. Analyses will be implemented natively in R 4.4.1 under Fedora Linux using a dependency-controlled renv environment. The complete analysis hierarchy is summarized in **Supplement 3.**

#### 2.11.2 Imaging Domain

The primary imaging estimand (**A1**) is the **within-reader ordinal shift in Q9 when 3DRA is added to DSA**. The unit of analysis is a first-pass patient × reader × phase rating. Q9 will be modeled using cumulative-link mixed-effects ordinal logistic regression with imaging phase as the fixed exposure of interest, reader as a fixed effect, and patient as a random intercept. The principal effect measure will be the common proportional odds ratio for assignment to a higher Q9 category under DSA+3DRA versus DSA alone. An odds ratio >1 therefore indicates a shift toward greater perceived implant adequacy after 3DRA is added.

For descriptive interpretation, patient–reader change will also be summarized as ΔQ9 = Q9_DSA+3DRA − Q9_DSA (range −4 to +4), with upward, unchanged, and downward transitions reported. The proportional-odds assumption will be evaluated; if materially violated, a less restrictive ordinal model will be used in a prespecified sensitivity analysis.

Secondary image-domain analyses will evaluate Q1–Q8 item-level changes, the number and direction of altered item assessments, absolute inter-reader agreement, agreement on the direction of change, intra-reader reliability in the concealed repeat subset, and Q10 technical image quality. Gwet AC2 with linear ordinal weights [48] will be the primary agreement/reliability coefficient; Krippendorff ordinal alpha and weighted Cohen kappa will be reported as supplementary measures. Agreement in absolute ratings is explicitly distinguished from agreement on change. Detailed A1–A5 specifications are provided in **Supplement 3**.

#### 2.11.3 Prognostic Domain

For prognostic analyses, the three independent first-pass Q9 ratings will be summarized at the patient level using the median under each imaging condition; with three readers, the median remains an observed ordinal category. The primary incremental exposure is the patient-level median change in Q9 between the DSA-only and DSA+3DRA conditions.

The prespecified clinical-control backbone (**M1**) contains age, sex, current symptomatic status, CFS ≥5, diabetes, contralateral carotid occlusion, significant baseline PHQ-2 screen, and stent architecture (double-layer vs single-layer). **M2** extends M1 by adding the patient-level median DSA Q9 assessment. **M3** then extends M2 by adding the patient-level median change in Q9 observed after 3DRA information becomes available. The principal prognostic comparison is **M2 versus M3**, which directly asks whether the change in expert assessment induced by access to 3DRA adds prognostic information beyond clinical-control information and conventional DSA. A secondary pragmatic model will use the final DSA+3DRA Q9 assessment directly as the full-information representation.

Primary multivariable predictive models will use ridge-penalized logistic regression to retain the locked predictors while limiting coefficient instability in the small cohort. The ridge penalty λ will be selected through outcome-stratified 5-fold cross-validation repeated 10 times, minimizing binomial deviance. Complementary Firth bias-reduced logistic regression [49] will address small-sample likelihood bias and potential separation. A prespecified parsimonious sensitivity family will assess robustness to a reduced clinical-control backbone under a frozen STOP rule that prohibits further variable deletion in response to observed model performance. Full model definitions are provided in **Supplement 3**.

#### 2.11.4 Internal Validation and Complementary Analyses

Prediction performance will be evaluated using discrimination (AUC), overall prediction error (Brier score), and calibration. Patient-level internal validation will use **2,000 bootstrap resamples**. For each ridge model, the entire tuning pipeline—including repeated cross-validation and λ selection—will be rerun within every bootstrap sample rather than treating the originally selected penalty as fixed. Optimism-corrected performance estimates for M2 and M3, together with paired incremental contrasts (ΔAUC and ΔBrier), will then be estimated.

Calibration assessment will include calibration-in-the-large/intercept, calibration slope, and graphical assessment. Because small-sample bootstrap correction of calibration slope can become numerically unstable, a prespecified reportability rule will determine whether an optimism-corrected slope can be interpreted. If stability criteria are triggered, the corrected slope will be retained in audit output but described as not reliably estimable rather than being trimmed, winsorized, or replaced by a more favorable estimator. Exact thresholds and sensitivity rules are provided in **Supplement 3**.

Secondary time-to-event analyses will include Kaplan-Meier estimates and ridge-penalized Cox models paralleling the principal model hierarchy, with RMST through day 104 as a prespecified sensitivity if substantial non-proportional hazards are supported. **Generalized Pairwise Comparisons for Trend (GPC-Trend)** will assess the association between ordered 3DRA-derived information and a prespecified patient-centered hierarchy spanning death, stroke, MI, END, major bleeding/surgical complication, additional unplanned revascularization, TIA, and readmission [50,51]. An exploratory **multi-reader multi-case (MRMC)** ROC analysis will evaluate whether access to 3DRA improves reader-level discrimination of subsequent TO failure using Q9 as the prespecified marker and the FDA iMRMC framework [52–54]. These analyses complement, but do not replace, A1 or the M2-versus-M3 comparison.

#### 2.11.5 Missing Data

Missingness will be summarized by variable and analysis domain. Clinical outcome events will not be imputed; participants lost to follow-up in time-to-event analyses will be censored at their last confirmed event-free assessment. Primary prognostic models will preferentially use observed complete predictor data.

Multiple imputation by chained equations will be used as a prespecified sensitivity analysis if **any primary predictor has >5% missingness or complete-case restriction removes >5% of the cohort**. The imputation model will include the corresponding analysis variables and relevant outcome information. Reader ratings will not be imputed for primary agreement analyses; paired phase-shift analyses will use observed paired ratings, with complete-pair sensitivity analysis where relevant. A Q10 rating of 1 represents poor technical evaluability rather than missingness and does not automatically exclude a case. The full sensitivity hierarchy is provided in **Supplement 3**.

## 3. Data Management and Open Science

### 3.1 Data Governance and Electronic Data Capture

Data stewardship is **FAIR**-aligned [55] while remaining subject to institutional governance and the Brazilian General Data Protection Law [19]. Direct identifiers are removed from analytical datasets and replaced by coded study identifiers. The re-identification key is stored separately on restricted-access encrypted institutional infrastructure at HCPA and is not included in public reproducibility materials.

Clinical, procedural, imaging-derived, and longitudinal outcome data are managed in REDCap [56] hosted on institutional infrastructure. Role-based access, structured fields, range and logic checks, mandatory-field constraints, audit trails, and institutional backup procedures support data integrity. Source information is retained separately from analysis-ready derivations so that analytical transformations do not silently overwrite adjudicated or source data.

### 3.2 Synthetic Data and Computational Reproducibility

A fully synthetic dataset will be generated from the locked analytical database using the synthpop R package [57] to support execution and verification of the analytical workflow when original participant-level data cannot be publicly released. The synthetic dataset is intended to preserve relevant marginal and multivariable structure sufficiently to support code execution and reproducibility testing; it is **not** intended to reproduce the original clinical estimates or substitute for participant-level data in independent clinical inference.

Before public release, synthetic-data utility and disclosure/re-identification risk will be evaluated to the extent technically feasible. The dataset will be explicitly labeled as simulated. Analytical code and reporting will preserve the distinction between validation of code behavior on synthetic data and scientific inference from the original locked dataset.

### 3.3 Reproducibility Repository and Controlled Data Access

After peer-reviewed publication of the primary study results, a version-controlled reproducibility package will be archived in a persistent repository such as the Open Science Framework with a static DOI. The package is intended to include data-cleaning and derivation code, reader-randomization documentation and locked seeds, statistical model code, bootstrap/internal-validation procedures, table/figure generation, the synthetic dataset, and the machine-readable study data dictionary.

Original participant-level data will remain restricted when public release is not permitted by consent, institutional governance, ethics approval, or Brazilian data-protection requirements. Qualified researchers may apply for controlled access through the HCPA Research Ethics Committee, subject to institutional and ethics approval. Public metadata, analysis code, and synthetic reproducibility materials are intended to remain accessible even when original participant-level data cannot be deposited without restriction.

## 4. Declarations and Administrative Information

### 4.1 Academic Framework and Acknowledgments

This investigation forms a core component of the doctoral thesis of Lucas Scotta Cabral in the Postgraduate Program in Cardiology and Cardiovascular Sciences (Programa de Pós-Graduação em Cardiologia e Ciências Cardiovasculares) at Universidade Federal do Rio Grande do Sul (UFRGS). The study is being conducted within the Division of Interventional Neuroradiology and the Cardiovascular Diagnosis and Therapy Unit at Hospital de Clínicas de Porto Alegre (HCPA), Porto Alegre, Brazil.

### 4.4 Ethical Approval, Consent, and Protocol Registration

The study protocol was approved by the Institutional Review Board / Research Ethics Committee at Hospital de Clínicas de Porto Alegre (Comitê de Ética em Pesquisa, CEP/HCPA; CAAE registration number: 76550023.4.3002.5330) and prospectively registered on the Registro Brasileiro de Ensaios Clínicos (REBEC: RBR-6cs4wcc). All participants or their legally authorized representatives provided written informed consent prior to enrollment and study procedures.

### 4.5 Author Contributions (CRediT Taxonomy)

- **Lucas Scotta Cabral (LSC)**: Conceptualization, Methodology, Software, Formal Analysis, Investigation, Data Curation, Writing – Original Draft, Visualization, Project Administration, Funding Acquisition.
- **Marco Vugman Wainstein (MVW)**: Conceptualization, Methodology, Supervision, Project Administration, Funding Acquisition, Writing – Review & Editing.
- **Sheila Cristina Ouriques Martins (SCOM)**: Methodology, Supervision, Project Administration, Writing – Review & Editing.
- **Rosane Brondani (RB)**: Validation, Investigation (Clinical Endpoint Adjudication), Writing – Review & Editing.
- **Juliana Ávila Duarte (JAD)**: Validation, Investigation (Neuroimaging Committee), Writing – Review & Editing.
- **Andrea Garcia de Almeida (AGA)**: Investigation (Transcranial Doppler Sub-study), Investigation, Writing – Review & Editing.
- **Angelica Dal Pizzol (ADP)**: Investigation (Transcranial Doppler Sub-study), Investigation, Writing – Review & Editing.
- **Raphael Machado de Castilhos (RMC)**: Methodology, Investigation (Cognitive & Functional Sub-study), Writing – Review & Editing.
- **Marco Antonio Stefani (MAS)**: Investigation (Brain Injury Biomarker Sub-study), Resources, Writing – Review & Editing.
- **SISYPHUS Study Executive Committee**: Investigation, Resources, Project Administration.

### 4.6 Data Availability Statement

Original participant-level data will not be made publicly available because of restrictions imposed by the approved consent/data-governance framework and Brazilian data-protection requirements (Lei Geral de Proteção de Dados, LGPD, Law No. 13.709/2018). Qualified researchers may apply for controlled access through the Research Ethics Committee of Hospital de Clínicas de Porto Alegre (Comitê de Ética em Pesquisa, CEP/HCPA; contact via), subject to institutional governance and ethics committee approval. To support computational reproducibility without compromising participant confidentiality, a fully synthetic research dataset generated with the synthpop package in R, accompanied by complete analysis code, locked random seeds, and data dictionaries, will be deposited in the Open Science Framework upon peer-reviewed publication.

## Study Registrations

REBEC (Registro Brasileiro de Ensaios Clínicos): RBR-6cs4wcc; Ethics Committee Approval (CAAE): 76550023.4.3002.5330 (HCPA); Protocol & SAP Linkage: Parent Protocol Version 4.0 (07/09/2026); Statistical Analysis Plan (SAP) Version 1.1.1 Clean (14/09/2026).

