## Supplement 1 for "SystematIc StudY of Patients with atHerosclerotic neUrovascular Stenoses (SISYPHUS): Study Protocol for a Prospective Observational Cohort Evaluating 3D Rotational Angiography and Final Implant Quality in Carotid Stenting"

### SISYPHUS FINAL IMPLANT QUALITY READER

### ASSESSMENT INSTRUMENT

#### S1.1. Purpose

This appendix reproduces the reader-facing Final Implant Quality assessment instrument used in the SISYPHUS image-reader study, together with an English publication translation. The original reader-facing wording is in Brazilian Portuguese. The English wording is provided for international reproducibility and does not replace the original instrument.

The instrument is administered within the standardized SISYPHUS image-reading environment. Readers are instructed to review each assigned case once within the designated reading session, using the permitted OHIF Viewer interactions, and to submit a single final set of Q1–Q10 ratings. Submitted ratings are locked and cannot be retroactively modified.

#### S1.2. Response scales

##### *Q1–Q8: Specific implant findings*

Five-category ordinal scale:

1. Definitely absent
2. Probably absent
3. Equivocal / indeterminate
4. Probably present
5. Definitely present

##### *Q9: Global implant adequacy*

Five-category ordinal scale:

1. Definitely inadequate
2. Probably inadequate
3. Equivocal / indeterminate
4. Probably adequate
5. Definitely adequate

##### *Q10: Technical image quality*

Three-category ordinal scale:

1. Poor / inadequate
2. Acceptable
3. Excellent

##### S1.3. Publication translation of the reader-facing instrument

| Item | Construct | English publication translation | Response scale |
|---|---|---|---|
| **Q1** | Stent malexpansion | What is the degree of suspicion/presence of **stent malexpansion** in the displayed images? | Q1–Q8 5-category scale |
| **Q2** | Significant mesh deformation or fracture | What is the degree of suspicion/presence of **significant stent-mesh deformation or fracture** in the displayed images? | Q1–Q8 5-category scale |
| **Q3** | Incomplete stent-wall apposition | What is the degree of suspicion/presence of **incomplete stent-mesh apposition (endoleak zones)** in the displayed images? | Q1–Q8 5-category scale |
| **Q4** | Eccentric plaque relative to the stent mesh | What is the degree of suspicion/presence of **plaque eccentric to the stent mesh** in the displayed images? | Q1–Q8 5-category scale |
| **Q5** | Residual plaque near or beyond stent margins | What is the degree of suspicion/presence of **residual plaque extending beyond the stent margins or located within 5 mm of a stent margin** in the displayed images? | Q1–Q8 5-category scale |
| **Q6** | Significant residual in-stent stenosis | What is the degree of suspicion/presence of **significant residual in-stent stenosis (>30%)** in the displayed images? | Q1–Q8 5-category scale |
| **Q7** | Mural thrombus | What is the degree of suspicion/presence of **mural thrombus** in the displayed images? | Q1–Q8 5-category scale |
| **Q8** | In-stent plaque protrusion | What is the degree of suspicion/presence of **in-stent plaque protrusion** in the displayed images? | Q1–Q8 5-category scale |
| **Q9** | Global implant adequacy | After evaluating the images for clinical decision-making, **how confident is the operator that the implant is adequate—that is, that the procedure has been completed successfully and satisfactorily?** | Q9 5-category adequacy scale |
| **Q10** | Technical image quality | What is the **technical image quality** for this case in the images evaluated? | Q10 3-category technical-quality scale |

#### S1.4. Interpretation

Q1–Q8 represent specific structural findings and are not assumed to carry equal clinical importance. No simple unweighted additive Q1–Q8 score is used as the primary Final Implant Quality measure.

Q9 is the primary integrated imaging variable. It captures the reader's global confidence that the carotid implant is satisfactory after integrating all visible stent–vessel–plaque information available in the assigned reading condition.

Q10 is a technical quality-control variable and is not incorporated into the Final Implant Quality construct.

#### S1.5. Original Brazilian Portuguese reader-facing wording

| Item | Original question in Brazilian Portuguese |
|---|---|
| **Q1** | Qual o grau de suspeição/presença do achado **"stent mal expandido"** na imagem demonstrada? |
| **Q2** | Qual o grau de suspeição/presença do achado **"deformidade significativa ou fratura de malha"** na imagem demonstrada? |
| **Q3** | Qual o grau de suspeição/presença do achado **"má aposição de malha (zonas de endoleak)"** na imagem demonstrada? |
| **Q4** | Qual o grau de suspeição/presença do achado **"placa excêntrica à malha"** na imagem demonstrada? |
| **Q5** | Qual o grau de suspeição/presença do achado **"placa residual além das margens do stent ou a menos de 5 mm das margens do stent"** na imagem demonstrada? |
| **Q6** | Qual o grau de suspeição/presença do achado **"existência de estenose residual significativa (>30%) intra-stent"** na imagem demonstrada? |
| **Q7** | Qual o grau de suspeição/presença do achado **"trombo mural"** na imagem demonstrada? |
| **Q8** | Qual o grau de suspeição/presença do achado **"protrusão de placa intra-stent"** na imagem demonstrada? |
| **Q9** | Ao avaliar a imagem e utilizá-la para tomada de decisão, qual **o grau de segurança do operador que o implante está adequado (ou seja, que foi finalizado com sucesso e de forma satisfatória)**? |
| **Q10** | Qual a **qualidade técnica da imagem** para este caso nas imagens analisadas? |

**S1.6. Reader-environment principles relevant to the instrument**

The instrument is completed within a standardized web-based, zero-footprint image-viewing environment. Readers are permitted to use the predefined core tools required for inspection of the frozen image package, including 2-dimensional navigation, pan, zoom, window/level adjustment, linear/angular measurement, series scrolling, and preset 3-dimensional volume-rendered rotations where available in the assigned package.

Advanced unconstrained external reprocessing, export of raw DICOM data to outside software, and reader-specific modification of the prepared package are not permitted.

Each case is assessed once per scheduled presentation. Ratings are finalized at the end of the reading session and locked after submission.

The reader-training cases are historical, non-study cases and do not enter any SISYPHUS analysis.
