## Supplement 2 for "SystematIc StudY of Patients with atHerosclerotic neUrovascular Stenoses (SISYPHUS): Study Protocol for a Prospective Observational Cohort Evaluating 3D Rotational Angiography and Final Implant Quality in Carotid Stenting"

### IMAGING ACQUISITION AND

### READER-STUDY IMPLEMENTATION

#### S2.1. Purpose

This appendix documents the operational implementation of the SISYPHUS imaging and image-reader study. It complements the main manuscript by specifying acquisition, package preparation, viewing, randomization, blinding, concealed repeat assessments, and reader-training procedures without changing the prespecified scientific estimands.

#### S2.2. End-of-procedure imaging acquisition

##### *S2.2.1. Completion digital subtraction angiography*

At the completion of carotid artery stenting (CAS), conventional completion digital subtraction angiography (DSA) is acquired in at least two orthogonal projections centered on the treated carotid segment. Both subtracted and non-subtracted runs are retained.

The completion DSA represents the baseline imaging information state used in Phase 1 of the reader study.

##### *S2.2.2. Three-dimensional rotational angiography*

Immediately after completion DSA, selective intra-arterial 3-dimensional rotational angiography (3DRA) is acquired without recrossing the freshly implanted stent with an additional intravascular imaging device.

The access catheter is positioned proximally to the stented segment. A standardized single-phase rotational acquisition is performed using high-definition reconstruction. A 10-mL aliquot of nonionic iodinated contrast is diluted with normal saline to a total volume of 50 mL (20% contrast concentration) and injected synchronously with the rotational acquisition.

The resulting volumetric dataset provides submillimeter 3-dimensional information for multiplanar and volume-rendered assessment.

##### *S2.2.3. Co-intervention control*

The investigational 3DRA reconstructions are not used to alter immediate procedural or postprocedural management. Participating interventionalists prospectively agreed not to use the experimental 3DRA interpretation for treatment modification, preserving separation between the investigational imaging exposure and subsequent clinical outcome ascertainment.

For prognostic analyses, completion of the 3DRA acquisition defines time zero.

#### S2.3. Image preparation and frozen information packages

Image preparation is standardized before formal reading. Reader packages are prepared independently of clinical outcome and are not modified according to reader identity, reader findings, or subsequent events.

The design principle is that every reader begins from the same prepared information set within each phase. Reader-dependent variation therefore occurs after exposure to a standardized package rather than through reader-specific source preparation.

#### S2.4. Viewing environment

Coded, de-identified DICOM datasets are centrally hosted on a secure Orthanc DICOM Server (version 1.12.3). Online case review and scoring are performed through a zero-footprint OHIF Viewer environment (version 3.8.0).

Readers may use the predefined core interactions required for clinical inspection:

- 2-dimensional navigation and series scrolling;
- pan and zoom;
- window/level adjustment;
- linear and angular measurements;
- inspection of the standardized prepared series;
- preset 3-dimensional volume-rendered rotations when included in the assigned package.

The following are prohibited:

- export of raw DICOM data to external software;
- unconstrained advanced external reprocessing;
- reader-specific reconstruction of an alternative package;
- custom algorithmic filtering outside the frozen workflow;
- post-submission modification of Q1–Q10 ratings.

#### S2.5. Readers, blinding, and reading conditions

Three board-certified interventional neuroradiologists, each with more than 5 years of post-specialization experience in vascular neurointerventions and neuroimaging interpretation, participate as readers.

Readers are independent of clinical endpoint adjudication and blinded to:

1. baseline clinical characteristics;
2. procedural details not contained in the standardized image package;
3. clinical outcomes;
4. the identity or repeat status of concealed repeat presentations.

Each reader evaluates every patient under two sequential, non-interleaved information conditions:

- **Phase 1 — DSA only:** the standardized completion-DSA package is reviewed without access to 3DRA.
- **Phase 2 — DSA + 3DRA:** the same patient's standardized DSA information is reviewed with the additional standardized 3DRA package.

The scientific estimand is therefore the effect of **access to additional 3DRA information under the prespecified sequential reading design**, not a randomized crossover comparison of modality order.

##### S2.6. Phase sequence and washout

All Phase 1 readings are completed before the same reader begins Phase 2. The phases are not interleaved.

A washout interval of at least 1 week separates completion of Phase 1 from initiation of Phase 2 for each reader.

Because DSA-only evaluation must precede exposure to 3DRA to preserve the baseline DSA information state, phase and presentation-order effects are intrinsically non-separable. Randomization is therefore applied to **case order within reader and phase**, not to modality sequence.

##### S2.7. First-pass and concealed repeat assessments

The first-pass reader-study volume is:

- 67 patients;
- 3 readers;
- 2 phases;
- 402 first-pass patient × reader × phase assessments.

A reproducibly randomized subset of 17 of 67 patients (25.4%) is used for concealed repeat presentation within each phase for each reader.

This contributes:

- 17 repeat patients × 3 readers × 2 phases = 102 concealed repeat assessments;
- **504 total reading assessments**.

Concealed repeats are used exclusively for intra-reader reliability and do not contribute additional observations to the primary DSA-versus-DSA+3DRA phase comparison.

#### S2.8. Blocking and randomized presentation order

Each reader-phase sequence contains 84 presentations, organized into five reading blocks:

- Block 1: 17 presentations;
- Block 2: 17 presentations;
- Block 3: 17 presentations;
- Block 4: 17 presentations;
- Block 5: 16 presentations.

Repeat presentations are placed in nonadjacent blocks to reduce overt recall.

Case order is independently randomized for each reader and each phase.

##### *S2.8.1. Locked random seeds*

| Sequence | Seed |
|---|---|
| Reader 1 — Phase 1 | 20261912 |
| Reader 1 — Phase 2 | 20261913 |
| Reader 2 — Phase 1 | 20261922 |
| Reader 2 — Phase 2 | 20261923 |
| Reader 3 — Phase 1 | 20261932 |
| Reader 3 — Phase 2 | 20261933 |

Repeat-subset seed: **20260911**.

Randomization code, seeds, internal patient-to-case mappings, repeat keys, and reader schedules are retained as provenance documentation.

##### S2.9. Rating workflow and locking

Each scheduled case presentation is reviewed in a single session. At completion of image inspection, the reader submits one final Q1–Q10 assessment.

Once submitted:

- the rating set is locked;
- the case cannot be reopened to revise the submitted score;
- prior ratings are not shown for comparison with subsequent cases;
- concealed repeat status remains hidden from the reader.

This single-submission workflow is intended to reduce retrospective score recalibration, anchoring to earlier cases, and post-hoc modification after later case exposure.

##### S2.10. Reader training

Before formal study readings, all readers complete a standardized training session using approximately 4–5 historical, non-study cases.

Training is designed to familiarize readers with:

- the OHIF workflow;
- permitted viewing tools;
- the Q1–Q10 scoring instrument;
- representative technical conditions and variation in image appearance.

Training is not used to calibrate readers toward a consensus answer. Training cases are excluded from every SISYPHUS analytical population.

##### S2.11. Analysis populations arising from the reader design

**Primary image-analysis population:** evaluable first-pass patient-reader-phase observations only.

**Inter-reader agreement population:** first-pass ratings within each imaging condition.

**Agreement-on-change population:** first-pass paired DSA and DSA+3DRA ratings.

**Intra-reader reliability population:** the concealed 17-patient repeat subset, analyzed separately by reader, phase, and item.

**Prognostic reader-derived exposure:** patient-level summaries use first-pass ratings only; concealed repeat ratings are excluded.

##### S2.12. Operational reproducibility

The reader-study implementation is preserved through:

- coded case identifiers;
- frozen package-preparation rules;
- standardized viewer and interaction set;
- locked randomization seeds;
- preserved reader-facing schedules;
- retained repeat-subset keys;
- version-controlled analysis scripts;
- reproducible documentation of first-pass and repeat-read populations.

These procedures are intended to permit reconstruction of the reading architecture without exposing patient identifiers or unblinding readers during study execution.
