## Supplement 3 for "SystematIc StudY of Patients with atHerosclerotic neUrovascular Stenoses (SISYPHUS): Study Protocol for a Prospective Observational Cohort Evaluating 3D Rotational Angiography and Final Implant Quality in Carotid Stenting"

### DETAILED STATISTICAL ANALYSIS FRAMEWORK AND PRESPECIFIED ANALYSIS HIERARCHY

#### S3.1. Purpose and interpretation hierarchy

This appendix summarizes the prespecified statistical architecture of SISYPHUS in a manuscript-oriented format. The governing Statistical Analysis Plan (SAP) remains the definitive source for implementation details and change control.

The formal interpretation hierarchy is:

1. **A1 — Primary imaging estimand:** within-reader ordinal shift in Q9 after addition of 3DRA.
2. **M2 versus M3 — Principal incremental prognostic comparison:** whether 3DRA-derived information adds prognostic information beyond the clinical-control backbone and DSA assessment.
3. All remaining analyses are secondary, sensitivity, or exploratory according to their prespecified role.

No secondary or exploratory result independently redefines study success.

#### S3.2. Analysis populations

*S3.2.1 Clinical analysis population*

All eligible participants in the locked prospective cohort (n=67).

S3.2.2 Primary image-analysis population

Evaluable first-pass patient-reader-phase observations. Concealed repeat presentations do not contribute additional observations to primary phase-comparison analyses.

*S3.2.3 Reliability population*

The reproducibly randomized 17-patient concealed repeat subset, analyzed separately for each reader and phase.

*S3.2.4 Unit of prognostic inference*

The patient is the unit of prognostic inference.

#### S3.3. Imaging-domain analyses

##### *S3.3.1. A1 — Primary within-reader ordinal Q9 shift*

Q9 is analyzed as a 5-category ordinal outcome using a cumulative-link mixed-effects ordinal logistic model:

$$logit\{P(Y_{ipr} \le k)\} = \theta_k - (\beta_{phase} X_{phase} + \beta_{reader}^{T} X_{reader} + u_i)$$

where:

- imaging phase is the fixed exposure of interest;
- reader is modeled as a fixed effect;
- patient is represented by a random intercept;
- only first-pass observations enter A1.

The principal effect measure is:

$$OR_{phase} = \exp(\beta_{phase})$$

where $OR_{phase} > 1$ indicates a shift toward greater perceived implant adequacy after addition of 3DRA.

##### *S3.3.2. Descriptive Q9 change*

For each first-pass patient-reader pair:

$$\Delta Q9 = Q9_{DSA+3DRA} - Q9_{DSA}$$

with possible values from −4 to +4.

The distribution of upward, unchanged, and downward shifts complements the primary ordinal model but does not replace it.

##### *S3.3.3. A2 — Secondary item-level change*

Q1–Q8 are analyzed as correlated secondary ordinal outcomes.

For item $j$:

$$\Delta Q_j = Q_{j,DSA+3DRA} - Q_{j,DSA}$$

For Q1–Q8, positive change indicates greater suspicion/presence of the abnormality after 3DRA. This direction differs from Q9, for which a positive shift indicates greater perceived adequacy.

The extent of changed assessment is summarized as:

$$N_{changed} = \sum_{j=1}^{8} I\left(Q_{j,DSA+3DRA} \neq Q_{j,DSA}\right)$$

*S3.3.4. A3 — Inter-reader agreement*

##### A3a: Absolute ratings

Agreement is evaluated separately under DSA and DSA+3DRA.

Primary coefficient:

- Gwet AC2 with linear ordinal weights.

Secondary/sensitivity coefficients:

- Krippendorff ordinal alpha;
- linearly weighted pairwise Cohen kappa.

##### A3b: Agreement on change

Each reader's first-pass change is reduced to:

$$D = \begin{cases} -1, & \Delta Q < 0 \\ 0, & \Delta Q = 0 \\ +1, & \Delta Q > 0 \end{cases}$$

Primary coefficient:

- Gwet AC2 with linear weights for the 3-level direction variable.

Complete 3-reader directional agreement is additionally summarized descriptively.

*S3.3.5. A4 — Intra-reader reliability*

The concealed 17-patient repeat subset is used exclusively for intra-reader reliability. Reliability is evaluated separately by reader, phase, and Q1–Q10 item.

Primary coefficient:

- Gwet AC2 with linear ordinal weights.

Supplementary measures:

- weighted Cohen kappa;
- exact agreement;
- agreement within ±1 ordinal category.

Repeat readings do not enter A1–A3 as additional observations.

*S3.3.6. A5 — Technical image quality and stent architecture*

Q10 is summarized separately as a technical quality-control variable.

The principal stent-architecture variable is:

- single-layer versus double-layer.

Exploratory analyses may assess:

- association between stent architecture and Q10;
- phase-by-stent-type interaction for Q9 and/or Q10.

These analyses do not alter A1.

#### S3.4. Patient-level representation of reader-derived imaging information

*S3.4.1. Primary representation*

For each patient:

$$Q9_{DSA}^{med} = median\left(Q9_{R1}, Q9_{R2}, Q9_{R3}\right)$$

$$Q9_{DSA+3DRA}^{med} = median\left(Q9_{R1}, Q9_{R2}, Q9_{R3}\right)$$

The incremental patient-level exposure is:

$$\Delta Q9^{med} = Q9_{DSA+3DRA}^{med} - Q9_{DSA}^{med}$$

*S3.4.2. Sensitivity representations*

Prespecified sensitivities include:

- arithmetic mean of the 3 first-pass readers;
- exploratory ordinal latent/empirical-Bayes representation;
- post-lock consensus only as an exploratory tool for discordant cases, never as the primary patient-level representation.

#### S3.5. Primary prognostic outcome

The principal prognostic outcome is 90-day Textbook Outcome (TO) failure.

For regression:

$$Y_{TO-failure} = \begin{cases} 1, & \text{failure to achieve TO} \\ 0, & \text{TO achieved} \end{cases}$$

The adjudicated endpoint is primary. Component-level reconstruction is retained for quality control and does not silently overwrite the adjudicated outcome.

#### S3.6. Clinical-control backbone

The full prespecified clinical-control model is:

**M1 =** Age + Sex + Current symptomatic status + CFS ≥5
+ Diabetes + Contralateral carotid occlusion
+ Significant baseline PHQ-2 screen + Stent architecture

No predictor is removed from the primary backbone because of univariable significance, automated selection, or variance-inflation-factor threshold alone.

Post-time-zero complications are not allowed to enter the primary clinical-control model.

#### S3.7. Locked prognostic model family

##### *S3.7.1. Full primary family*

$$M_2 = M_1 + Q9^{med}_{DSA}$$

$$M_3 = M_2 + \Delta Q9^{med}$$

$$M_{3R} = M_1 + Q9^{med}_{DSA+3DRA}$$

| Model | Information state | Scientific role |
|---|---|---|
| **M1** | Clinical-control information | Baseline prognostic control |
| **M2** | M1 + DSA Q9 | Prognosis after adding conventional DSA-derived global assessment |
| **M3** | M2 + ΔQ9 | **Primary extended model: incremental 3DRA information beyond DSA** |
| **M3R** | M1 + final DSA+3DRA Q9 | Pragmatic full-information replacement model |

The principal prognostic comparison is **M2 versus M3**.

##### *S3.7.2. Parsimonious sensitivity family*

The locked parsimonious backbone is:

**M1P =** Age + Current symptomatic status + Diabetes + Contralateral carotid occlusion + Stent architecture

with:

$$M_{2P} = M_{1P} + Q9^{med}_{DSA}$$

$$M_{3P} = M_{2P} + \Delta Q9^{med}$$

$$M_{3RP} = M_{1P} + Q9^{med}_{DSA+3DRA}$$

**STOP RULE:** no further covariate deletion is permitted in response to real-data AUC, Brier score, calibration, p values, coefficient direction, or imaging effects.

#### S3.8. Primary prognostic estimation strategy

##### *S3.8.1. Ridge-penalized logistic regression*

The principal predictive framework uses ridge-penalized logistic regression with an L2 penalty.

The penalty parameter $\lambda$ is selected using outcome-stratified 5-fold cross-validation repeated 10 times, minimizing mean binomial deviance.

The complete tuning procedure is repeated inside every bootstrap resample.

##### *S3.8.2. Firth logistic regression*

Firth bias-reduced logistic regression is the principal complementary inferential/sparse-data approach.

It is used to provide interpretable association estimates and profile penalized-likelihood confidence intervals where estimable.

Firth modeling complements but does not replace the ridge prediction pipeline.

##### *S3.8.3. Conventional maximum-likelihood logistic regression*

Conventional logistic regression is supplementary when estimable and does not govern interpretation in the presence of separation or severe sparse-data instability.

##### *S3.8.4. LASSO*

LASSO is **not** part of the primary prognostic architecture because automatic deletion of prespecified clinical-control variables conflicts with the scientific objective of retaining the locked backbone.

#### S3.9. Prognostic performance

No single statistic defines model performance.

##### *S3.9.1. Discrimination*

Primary discrimination metric:

- C-statistic / AUC.

Principal incremental contrast:

$$\Delta AUC = AUC_{M3} - AUC_{M2}$$

##### *S3.9.2. Overall prediction error*

Primary overall prediction-error metric:

- Brier score.

Principal incremental contrast:

$$\Delta BS = BS_{M3} - BS_{M2}$$

Negative ΔBrier favors M3.

##### *S3.9.3. Calibration*

Calibration assessment includes:

- apparent calibration intercept/calibration-in-the-large;
- apparent calibration slope;
- calibration plot;
- classical 2,000-bootstrap mean-optimism correction where numerically interpretable.

##### *S3.9.4. Calibration-slope reportability rule*

The classical mean-optimism-corrected calibration slope is classified as unstable for primary interpretation if any of the following occurs:

1. 10% of test-on-original calibration slopes are non-positive;
2. 5% have absolute test-on-original slope >10;
3. 5% of test-on-original linear predictors have SD <0.05;
4. absolute difference >2 between classical mean-optimism and median-optimism corrected slopes;
5. absolute classical mean-optimism corrected slope >10.

If any criterion is triggered:

- the corrected value remains preserved in machine-readable audit output;
- it is not interpreted as a valid primary corrected slope;
- the manuscript reports that the corrected slope was **not reliably estimable by mean-optimism bootstrap under the prespecified stability rule**;
- apparent calibration and other performance measures remain reportable.

Extreme bootstrap replicates are not deleted or winsorized merely to obtain a more favorable calibration estimate.

##### S3.10. Bootstrap internal validation

Patient-level internal validation uses **2,000 bootstrap resamples**.

Within each bootstrap replicate:

1. 67 patients are sampled with replacement;
2. the complete prespecified model is rebuilt;
3. ridge penalty tuning is repeated within the bootstrap sample;
4. repeated outcome-stratified 5-fold cross-validation is rerun;
5. duplicate draws of an original patient remain in the same internal tuning fold to avoid leakage;
6. apparent bootstrap performance is calculated;
7. the bootstrap-fitted model is applied to the original cohort;
8. optimism is calculated as bootstrap apparent minus test-on-original performance;
9. optimism correction is applied where numerically interpretable.

Incremental M2-versus-M3 contrasts are corrected within paired bootstrap replicates.

This procedure constitutes internal validation and is not described as external validation.

#### S3.11. Time-to-event secondary analyses

##### *S3.11.1. Time origin and censoring*

Time zero is completion of the 3DRA acquisition.

General time-to-event censoring is:

$$T_i = min\left(T_{first qualifying event}, T_{last confirmed event-free follow-up}, 104\right)$$

Component-specific windows remain in force:

- END: ≤48 hours;
- BARC ≥3 / Clavien-Dindo ≥3: through discharge, maximum 72 hours;
- death, stroke, MI, additional unplanned revascularization, TIA, and unplanned readmission: through last valid follow-up, capped at day 104.

##### *S3.11.2. Kaplan-Meier and Cox framework*

Where informative:

- Kaplan-Meier estimates of freedom from first TO-defining event;
- ridge-penalized Cox models paralleling M1/M2/M3/M3R;
- Efron tie handling for primary ridge/conventional Cox models.

##### *S3.11.3. Proportional-hazards contingency*

Proportional hazards are assessed using Schoenfeld-residual diagnostics and graphical review.

If substantial non-proportional hazards are supported, restricted mean event-free time through $\tau = 104$ days is evaluated as a prespecified sensitivity measure.

#### S3.12. Generalized pairwise comparisons for trend

*S3.12.1. Role*

GPC-Trend / win-statistics analyses are complementary secondary analyses and do not replace the primary binary TO model or M2-versus-M3 comparison.

Primary ordered imaging exposure:

$$\Delta Q9^{med}$$

Secondary ordered exposure:

$$Q9^{med}_{DSA+3DRA}$$

Principal association measure:

generalized odds ratio for trend (GOR-trend),

with 95% patient-bootstrap percentile CI.

*S3.12.2 Locked clinical hierarchy for win statistics*

Death > Stroke > MI > END >
Major bleeding/surgical complication >
Any additional unplanned revascularization >
TIA > Any unplanned readmission

*S3.12.3. Pairwise rules*

For each patient pair with unequal ordered exposure:

- neither has the event at a hierarchy level → tie and proceed;
- one has the event → the event-free patient wins;
- both have the event → the patient with the later event wins;
- identical observed event time → tie and proceed.

The major bleeding/surgical-complication level is treated as a binary presence/absence construct for GPC. The patient remains the unit of inference. Pairwise comparisons are not treated as independent observations.

*S3.12.4. Reporting*

Where estimable, reporting includes:

- GOR-trend;
- 95% patient-bootstrap percentile CI;
- exploratory 2-sided p value;
- win/loss/tie probabilities;
- net benefit;
- win odds / win ratio;
- exposure-tied pairs separately;
- contribution of each clinical hierarchy level to resolved wins/losses.

#### S3.13. Exploratory MRMC prognostic discrimination

*S3.13.1. Scientific question*

Does access to DSA+3DRA improve individual-reader discrimination of subsequent 90-day TO failure compared with DSA alone?

*S3.13.2. Analysis population*

First-pass observations only:

- 67 patients;
- 3 readers;
- 2 conditions;
- 402 first-pass ratings.

Concealed repeat readings are excluded.

*S3.13.3. Marker and orientation*

Q9 is the only MRMC marker.

Because higher Q9 indicates greater adequacy while TO failure is adverse:

$$Q9_{risk} = 6 - Q9$$

*S3.13.4. Principal estimand*

$$\Delta AUC_{MRMC} = \overline{AUC}_{DSA+3DRA} - \overline{AUC}_{DSA}$$

*S3.13.5. Inference framework*

Primary exploratory inference:

- FDA iMRMC U-statistic random-reader/random-case framework.

Cross-check / sensitivity:

- Obuchowski-Rockette/Hillis random-reader/random-case analysis;
- fixed-reader/random-case sensitivity.

Three readers limit precision of reader-population variance estimation; numerical limitations are reported transparently rather than silently replaced.

*S3.13.6. MRMC non-goals*

The prespecified MRMC analysis does **not** include:

- M1 clinical covariates;
- MRMC comparison of M2 versus M3;
- reader-specific multivariable prognostic models;
- separate Q1–Q8 MRMC batteries;
- data-driven selection of a best reader;
- consensus Q9 as ground truth;
- primary dichotomization of Q9;
- alternate marker selection after observing AUC results.

#### S3.14. Missing data

*S3.14.1. Clinical outcomes*

Clinical outcome events are not imputed.

Loss to follow-up in time-to-event analyses results in censoring at the last confirmed event-free assessment.

*S3.14.2. Primary predictors*

Observed complete predictor data are preferred.

MICE is triggered as a prespecified sensitivity analysis if either:

- any primary predictor has >5% missingness; or
- complete-case restriction removes >5% of the clinical cohort.

*S3.14.3. Reader ratings*

Missing reader ratings are not imputed for primary agreement analyses.

Primary phase-shift sensitivity analyses may be restricted to complete patient-reader pairs.

Q10=1 (poor/inadequate technical image quality) is a real evaluability state and is not treated as missingness or an automatic exclusion.

**S3.15. Sensitivity-analysis hierarchy**

| Domain | Prespecified sensitivity / complementary analysis | Role |
|---|---|---|
| Imaging | Less restrictive ordinal model if proportional odds materially violated | Sensitivity |
| Imaging | Reader-by-phase interaction | Secondary |
| Imaging | Mean rather than median patient-level Q9 | Sensitivity |
| Imaging | Exploratory latent/empirical-Bayes Q9 | Exploratory |
| Agreement | Alternative agreement coefficients | Sensitivity |
| Prognosis | Parsimonious M1P/M2P/M3P/M3RP family | Sensitivity |
| Prognosis | Firth logistic regression | Complementary |
| Prognosis | Conventional MLE logistic regression | Sensitivity |
| Prognosis | Adverse Outcome instead of TO | Sensitivity |
| Survival | Ridge Cox / parsimonious Cox | Secondary / sensitivity |
| Survival | Firth Cox | Complementary |
| Survival | RMST if substantial non-proportional hazards | Sensitivity |
| Missing data | MICE if prespecified trigger reached | Sensitivity |
| MRMC | OR/Hillis random-reader/random-case cross-check | Exploratory cross-check |
| MRMC | Fixed-reader/random-case analysis | Exploratory sensitivity |

**S3.16. Multiplicity and interpretation**

Multiplicity is addressed principally through hierarchy rather than a family-wise confirmatory correction across every secondary analysis.

The study has:

- one primary imaging estimand (A1);
- one principal incremental prognostic comparison (M2 versus M3).

Secondary and exploratory *p* values cannot independently redefine the primary study conclusion.

Interpretation emphasizes effect magnitude, direction, precision, calibration, internal consistency, and the limitations imposed by the effective sample size.

**S3.17. Reproducibility**

Primary analyses are implemented in R under a frozen dependency-controlled environment.

The reproducibility record includes:

- R and package versions;
- operating environment;
- random-number seeds;
- dependency lock file;
- reader-randomization code;
- derivation scripts;
- model-fitting code;
- bootstrap-validation code;
- table/figure generation code;
- machine-readable outputs and quality-control logs.

Dummy/synthetic data may be used to validate code behavior and reporting logic but have no scientific inferential role.
